# A Cost-effectiveness analysis of Nivolumab plus chemotherapy for the first-line treatment of locally advanced or metastatic gastric/GEJ/oesophageal adenocarcinoma in the United States of America

**DOI:** 10.1101/2024.02.23.24303251

**Authors:** Jin Zhou, Yukai Tang, Geli Li

**Author notes:** 1st set of equal contributors (Jin Zhou, Yukai Tang).

## Abstract

**Background:** Nivolumab in combination with chemotherapy significantly improves survival in patients with gastric/gastroesophageal junction (G/GEJ)/esophageal adenocarcinoma.The purpose of this study was to evaluate the cost-effectiveness of Nivolumab plus chemotherapy for G/GEJ/esophageal adenocarcinoma.

**Methods:** A Markov model was developed on the basis of the US healthcare payers’ perspectives. We estimated the costs and summarised their effectiveness as quality-adjusted life-years (QALYs). One-way and probabilistic sensitivity analyses were conducted to explore the impact of uncertainties on the cost-effectiveness’s results.

**Results:** The incremental cost-effectiveness ratios (ICER) for Nivolumab plus chemotherapy($149636.97,1.24QALYs) verus chemotherapy($13941.06,0.75QALYs) is $135695.91 and the QALYs is 0.49.

**Conclusions:** Evidence suggests that Nivolumab plus chemotherapy a for the first-line treatment of locally advanced or metastatic gastric/GEJ/oesophageal adenocarcinoma may be not a cost-effective choice.

## 1. Background

Gastric/gastroesophageal junction(G/GEJ)/oesophageal adenocarcinoma can be regarded as a relatively common cancer worldwide, and its incidence is increasing day by day. Nowadays, gastric cancer is the fifth most common cancer in the world, and its fatality rate ranks the third among all cancers. From the perspective of geographical distribution, the incidence has increased significantly in east Asia in recent years, as has North America (1). At present, surgery is still the preferred treatment for G/GEJ/oesophageal adenocarcinoma. However, the prognosis is bleak and the postoperative recurrence rate reaches 50%-70% (2, 3). For those patients with unresectable advanced stage, other methods can only be sought.

In recent years, many studies have focused on improving the prognosis of this group of patients. At present, fluorouracil, platinum and paclitaxel are the main chemotherapy drugs for advanced G/GEJ/oesophageal adenocarcinoma with negative HER-2, but their therapeutic effect is not optimistic, and the survival time of most patients receiving chemotherapy is not more than 1 year (4–6). Besides, after the use of targeted therapy combined with chemotherapy for these patients, their survival was not significantly improved compared with chemotherapy (7–10).

Nivolumab is a PD-1 inhibitor. As PD-L1 is expressed more obviously in tumor cells and tumor immune cells than in normal cells, PD-1 inhibitor has been gradually promoted to a higher position in tumor therapy. Nivolumab has shown good efficacy in the treatment of many cancers such as non-small cell lung cancer and renal cell carcinoma (11, 12). Therefore, the Checkmate 649, a phase 3 trial, has been carried out,in which researchers recruited a group of advanced, unresectable and HER-2 negative patients and randomly divided them into two groups. In addition to the analysis of all patients, PD-L1 CPS≥5 patients were singled out for follow-up analysis. Then, the prognosis of the two groups of patients was compared between the combination chemotherapy with Nivolumab and chemotherapy alone.In the Result, For patients with PD-L1 CPS≥5, median OS improvement was 3 months (14.4 months [95% CI 13.1 to 16.2]vs 11.1 month [10.0 to 12.1]); Median PFS was 7.7 months (95% CI 7.0 to 9.2) for combination chemotherapy with Nivolumab compared with 6.05 months (5.0 to 6.9) for chemotherapy. For all randomized patients, the median OS was 13.1 months (IQR, 6.7 to 19.1) in the Nivolumab group and 11.1 months (5.8 to 16.1) in the chemotherapy group. In conclusion, Compared with chemotherapy alone, Nivolumab combined with chemotherapy significantly improved patient survival, especially in patients with PD-L1 CPS≥5 (13).

A cost-benefit analysis is necessary before a new drug becomes an approved method of treatment for many patients. Hence, in this study, based on the results of the Checkmate649 clinical trial, we established an economic model to evaluate the cost-effectiveness of Nivolumab in combination with chemotherapy in G/GEJ/oesophageal adenocarcinoma.

## 2. Materials and Methods

### 2.1 Population

CheckMate 649 is a randomized phase 3 trial involving 1581 patients from From March 2017 through April 2019 in all over the world. Median progression-free survival (7·7 months [95% CI 7·0–9·2]) was significantly longer in the nivolumab plus chemotherapy group than in the chemotherapy group (6.05 months [95% CI 5.6–6.9]) in patients with PD-L1 CPS ≥5, as was median overall survival analysis (14·4 months [95% CI 13·1–16·2] *vs* 11·1 months [10·0–12·1], respectively) in patients with PD-L1 CPS ≥5).Our data were based on clinical characteristics of CheckMate 649 subjects aged 18 years or older with previously untreated, unresectable advanced or metastatic gastric, GEJ, or oesophageal adenocarcinoma, regardless of PD-L1 expression.

We focused on the third phase of the CheckMate 649 study. In the total number of people in this trial, there are 955 patients with PD-L1 CPS≥5. Among them, 473 participants were in the experimental group(Nivolumab plus chemotherapy)and 482 in the control group(chemotherapy).We conducted a cost-effectiveness analysis for the Nivolumab plus chemotherapy group and chemotherapy group to provide a foundation for their different treatment. The research methods refer to the consolidated health economic evaluation reporting standards (CHEERS)( see Supplementary Information S1).

### 2.2 The Model’s Structure

The study used TreeAge Software 2021 (TreeAge Software, Inc, Williamstown, Massachusetts) to programme a multi-state Markov model. The purpose was to evaluate the cost-effectiveness of Nivolumab plus chemotherapy and chemotherapy in patients with untreated, unresectable advanced or metastatic gastric, GEJ, or oesophageal adenocarcinoma.This was due to the Markov model being able to provide more flexible modelling assumptions.(14) The multiple health states include PFS, progressive disease state (PD) and death. Assuming that patients in a certain state only make one state transition in a cycle, once the patients are in the PD state, they cannot return to the PFS state. Similarly, the patients in the dead state cannot transition to other states. The specific transition relationships are shown in Figure 1. We assumed that all the patients were in a PFS healthy state at the model’s initial stage. The patients were treated with Nivolumab plus chemotherapy or chemotherapy according to their groupings. When the disease progresses, the follow-up treatment plan in the CheckMate 649 clinical trial is used for treatment until the patient’s death.

**Figure 1.**
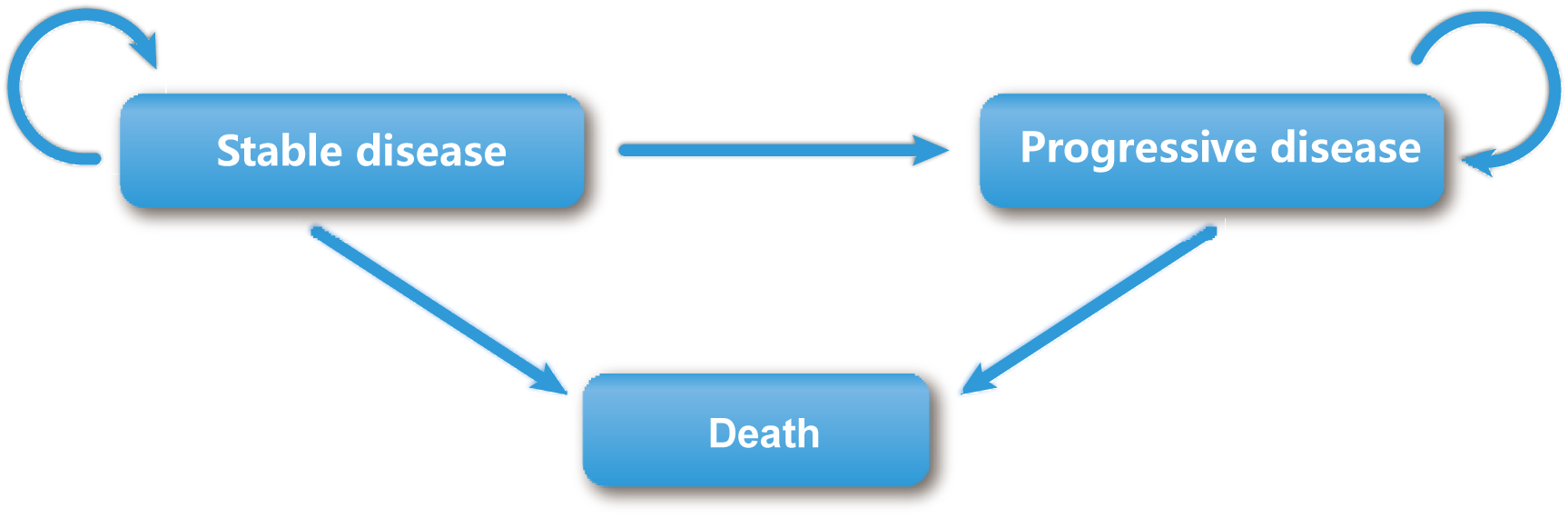
Markov state transition model.

We developed a Markov model to simulate the patient’s entire life course and evaluate the cost and effectiveness of first-line therapy for patients with advanced/metastatic gastric, GEJ, or oesophageal adenocarcinoma.In the CheckMate 649 clinical trial, the median survival time of the experimental group was 14.4 months, the control group was 11.1 months, and the overall time of the study is approximately 2 years. The effect of immunotherapy has a delayed effect and may continue to work beyond the treatment period. It should be analyzed from long-term data to avoid inaccuracy and uncertainty of results(15, 16).As the result of that,with reference to the dosing cycle of the CheckMate 649 clinical trial, we set the cycle of the Markov model to three weeks and the time range was 10 years. Approximately 99% of patients were in the absorption state.(17) A half-circle correction was conducted to simulate the transfer process more accurately. Simultaneous simulation analysis of the cost and utility is performed to estimate the cumulative total cost and health outcomes within the cohort’s time frame.(18, 19) The research was based on the American payers’ perspectives, with a 3% discount on costs and utilities.(20) According to the World Health Organization, ICER is acceptable when it is below three times GDP per capita(21). This study will use three times of the United States’s triple GDP per capita in 2021($69219.5) as the threshold(https://fred.stlouisfed.org/series/A939RC0Q052SBEA).The WTP is assumed to be $207659. The research indicators include the costs, life-years (LYs), quality-adjusted life-years(QALYs), and the incremental cost-effectiveness ratios (ICERs).

### 2.3 The Model’s Survival and Progression Risk Estimates

The original data for constructing the model were obtained from the CheckMate 649 clinical trial. When some data were unavailable, we referred to the related published literature. The GetData Graph Digitiser (version 2.26; http://getdata-graph-digitizer.com/download.php) was used to extract the Kaplan–Meier curve’s data of the PFS and OS in the Nivolumab plus chemotherapy group and the chemotherapy group. We also referred to the algorithm of Guyot et al. who refers to the pseudo-individual patient’s data reconstructed by R software (version 4.1.0; https://www.r-project.org/).(22) This was combined with the Akaike Information Criterion (AIC) and the Bayesian Information Criterion(BIC) to select the Log-logistic distribution that fitted the survival curve for Nivolumab plus chemotherapy and chemotherapy respectively after the reconstruction(eTable 1).(23) The distribution has a higher flexibility and estimated correlations.(24, 25)

### 2.4 The Utility and Cost Estimates

During the follow-up, the CheckMate 649 trial used the Gastric Cancer Subscale (GaCS) to compare quality of life after Nivolumab plus chemotherapy vs chemotherapy as the first-line treatment for advanced gastric cancer/gastroesophageal junction cancer/oesophageal adenocarcinoma.the average health utility (0.797 for PFS and 0.577 for PD) of the patients with gastric cancer/gastroesophageal junction cancer/oesophageal adenocarcinoma in the PFS and PD was obtained by previously published study(26). The top three incidence adverse events(AEs) with grade 3 or above were selected in Nivolumab plus chemotherapy (nivolumab [360 mg every 3 weeks or 240 mg every 2 weeks]-pluschemotherapy[XELOX every 3 weeks or FOLFOX every 2 weeks], nivolumab-plus-ipilimumab) and chemotherapy to be considered to evaluate the loss of the health utility caused by the three to five adverse events(AEs) for simplifying the calculation.The top three incidence adverse events(AEs) with grade 3 or above in Nivolumab plus chemotherapy and chemotherapy are Neutropenia(14%),Decreased platelet count(8%),Proteinuria(5%),Increased blood bilirubin(5%), Increased γ-glutamyltransferase(5%) and Palmar-plantar erythrodysaesthesia syndrome(12%), Hypertension(6%), Increased aspartate aminotransferase (5%), respectively.

The costs are reported in 2021 US dollars (US $1.0 = CNY Ɏ6.38).Only the direct costs of the medical expenses were considered. This included the cost of the drugs, subsequent treatment costs, management costs, follow-up costs, laboratory examination costs, and the major adverse reactions with grade 3 or above had the top three incidence rates according to CheckMate 649 trial.

The drug prices we selected local hospital prices or the price obtained by consulting with the drug supplier. The estimated cost of each drug during the set period is listed in Table 2. The probability that different treatment groups intend to receive different follow-up treatment and the treatment mode of specific subsequent therapies(systemic therapy other than PD-(L)1 inhibitors, local regional therapy, radiation therapy, surgery, PD-(L)1 inhibitors) are derived from CheckMate 649 trial.

**Table 1.**
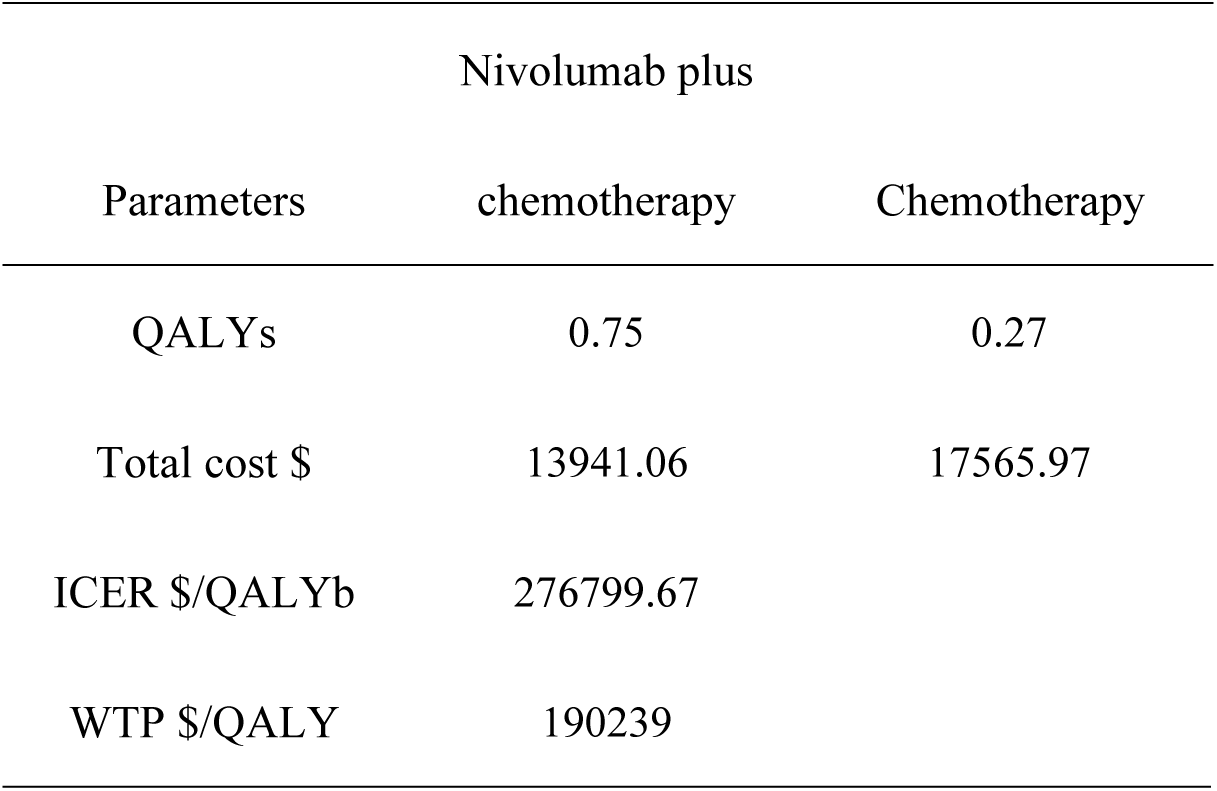
Base-case Analysis.

**Table 2.**
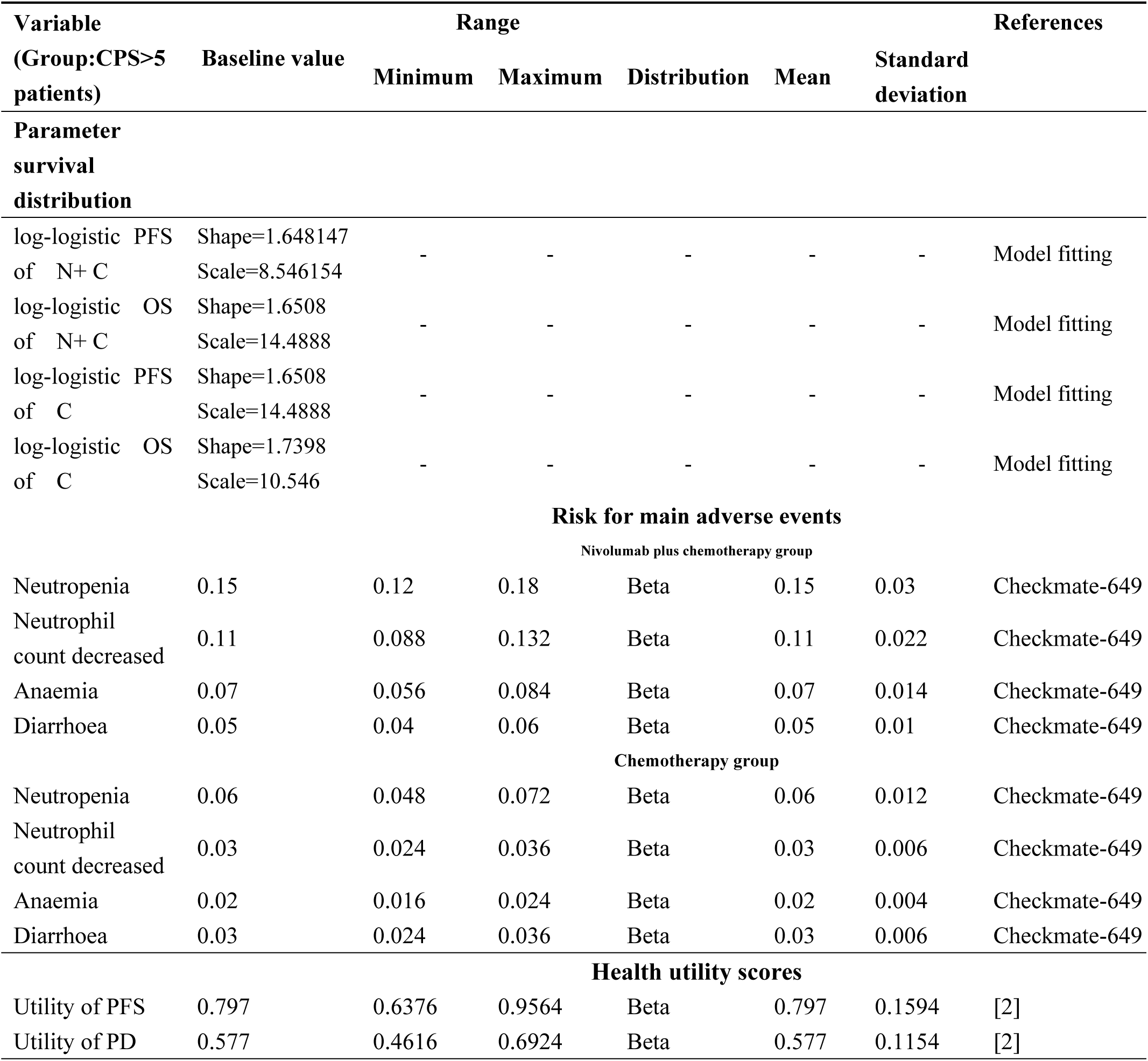
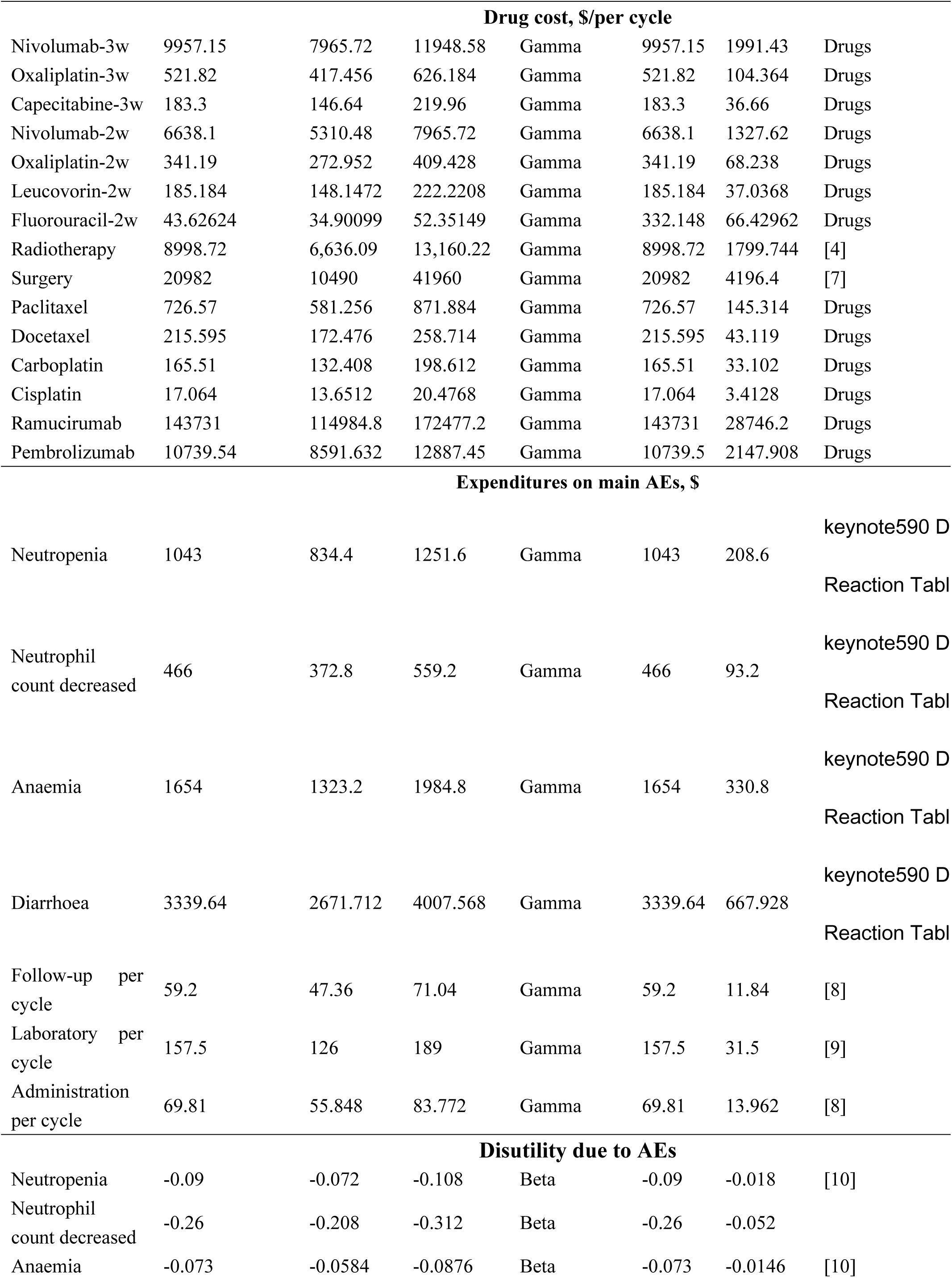
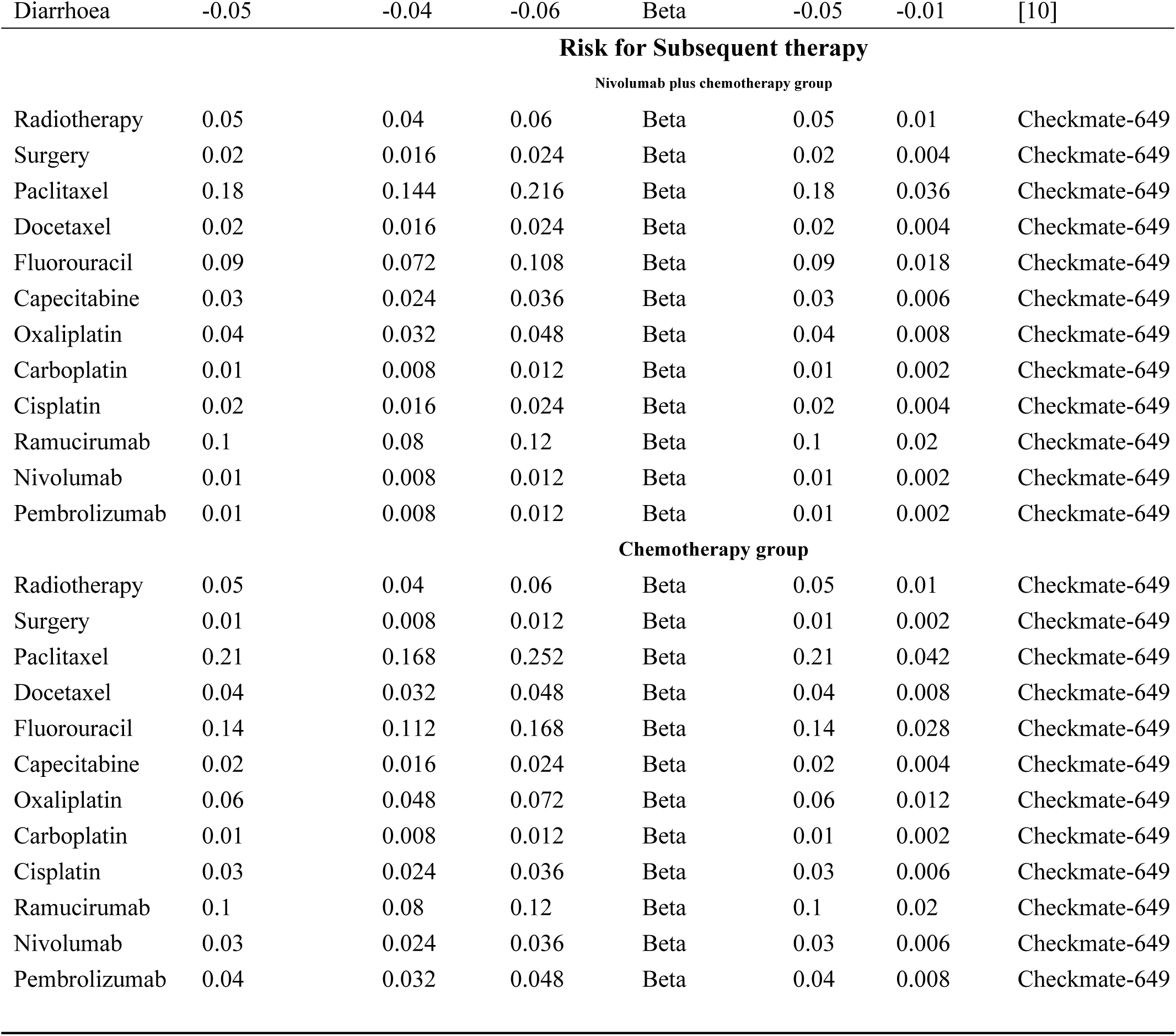
Model parameters: baseline values, ranges, and distributions for sensitivity analysis. Main model parameters.

The calculated drug dose are based on the actual clinical trials. In the Nivolumab plus chemotherapy group, the patients received 360 mg of Nivolumab every 3 weeks or 240 mg every 2 weeks and chemotherapy (XELOX [capecitabine 1000 mg/m2 twice daily, days 1–14 and oxaliplatin 130 mg/m^2^, day 1, every 3 weeks] or FOLFOX [leucovorin 400 mg/m2, day 1, fluorouracil 400 mg/m^2^, day 1 and 1200 mg/m2, days 1–2, and oxaliplatin 85 mg/m2, day 1, every 2 weeks]). In the chemotherapy group, the patients received chemotherapy alone. We assumed that the average body surface area was 1.68 m^2^.(27)When patients disease progressed, we assumed that all patients who disease progressed had follow-up treatment. It is important to note that the systemic therapy other than PD-(L)1 inhibitors in the subsequent therapies for advanced Gastric cancer, which we chose based on the NCCN 2022.1 guideline, is oxaliplatin, leucovorin plus fluorouracil therapy(oxaliplatin 85 mg/m2 IV on day 1, Leucovorin 200 mg/m2 IV on day 1,2,fluorouracil 2600 mg/m2 IV continuous infusionover 24 hours on day1).

### 2.5 Sensitivity Analyses

A one-way sensitivity analysis was performed to explore the influence of uncertain parameters on the ICER.

Each parameter was independently changed by assuming ±20% of the expected value to determine the obvious influence on decision-making.

Probabilistic analysis (PSA) was used to randomly sample all the parameters from a specified distribution to further explore the uncertainty and relevance of the model’s parameters. According to the parameter type, we selected the appropriate distribution for each uncertain parameter: the cost of the adverse reactions to drugs and treatment is the gamma distribution. The risk of adverse reactions, and the health utility scores including PFS, OS, and AE are the beta distribution. We performed a second-order Monte Carlo simulation of 10,000 iterations and generated a cost-benefit acceptability curve (CEAC) to show that Nivolumab plus chemotherapy is cost-effective with different WTP thresholds.

## 3. Results

### 3.1 Base-case Analysis

The result of base-case analysis about the cost and effectiveness of Nivolumab plus chemotherapy group and chemotherapy group in patients with untreated, unresectable advanced or metastatic gastric, GEJ, or oesophageal adenocarcinoma was shown in Table 1.According to our analysis,the incremental cost of Nivolumab plus chemotherapy($149636.97,1.24QALYs) verus chemotherapy($13941.06,0.75QALYs) is $135695.91 and the QALYs is 0.49. The ICER values($276799.67) are higher than the United States’s triple GDP per capita threshold in 2021 ($207659).

### 3.2 Sensitivity Analyses

A one-way sensitivity analysis was used to test the robustness of the two population model outputs. Under the condition that the input model parameters change by ±20%,the influence of each parameter on the analysis results is explored.The results are presented in the tornado diagram (Figure 2).The sensitivity analysis results demonstrated that the cost of Nivolumab has the most contributed to it.

**Figure 2.**
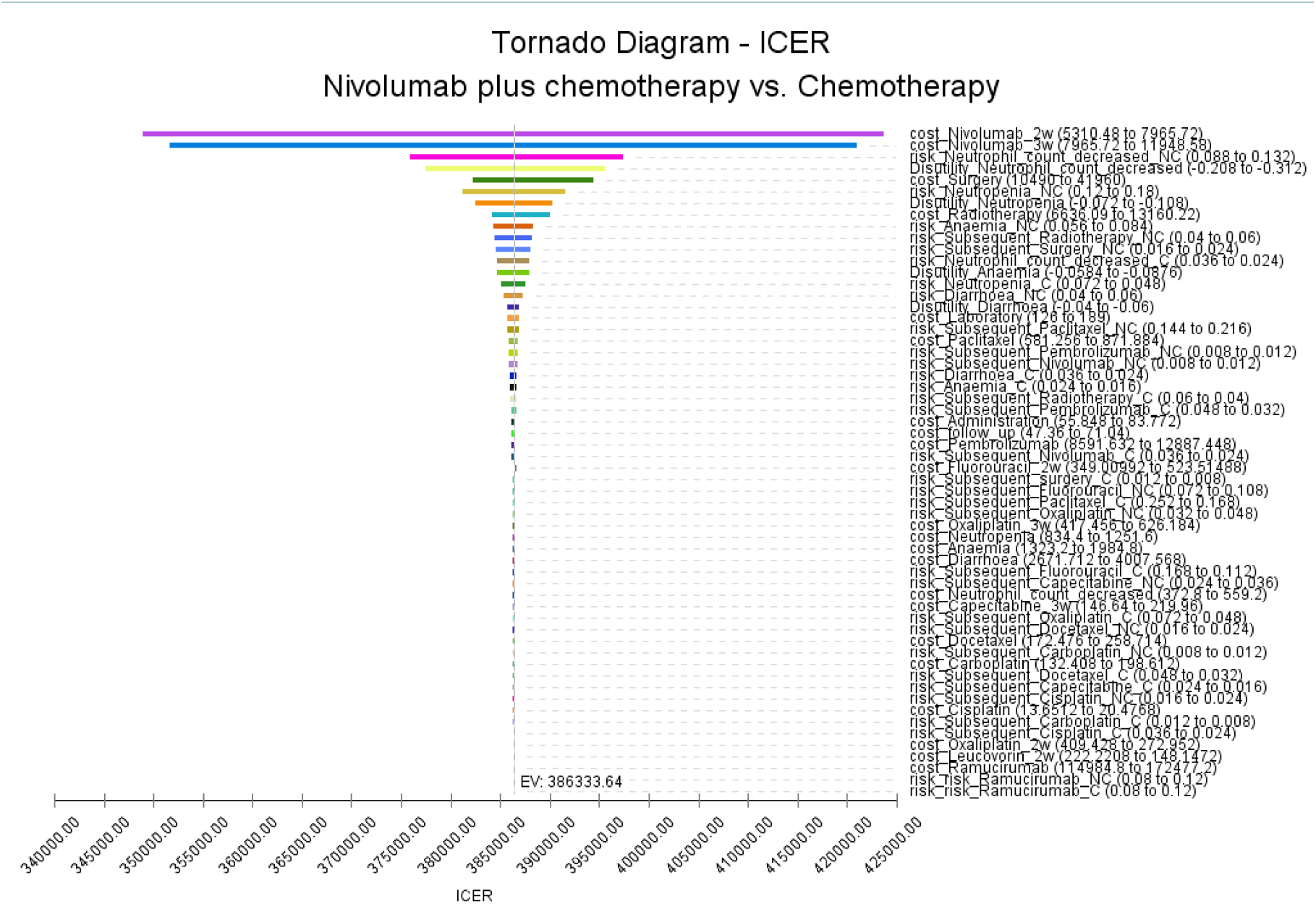
Tornado diagram for one-way sensitivity analysis. Abbreviation: SD= Stable disease; PD= Disease progression; ICER= Incremental cost-effectiveness ratio.

### 3.3 Probability sensitivity analysis

Probability sensitivity analysis (PSA) is applied to test the bias of the multiple model parameters on the analysis results when the multiple model parameters change simultaneously. The results are presented through cost-effectiveness acceptability curves (Figure 3) and incremental cost-effectiveness scatterplots (Figure 4). According to the results, we found that the higher the average social willingness to pay, the higher the probability of Nivolumab plus chemotherapy producing the cost effect.Under the condition of a payment threshold of $207659 per QALY, Probabilistic sensitivity analysis showed that there was 1.49% probability that Nivolumab plus chemotherapy was cost-effective within the fluctuation range of other model parameters in first-line in advanced gastric, GEJ, or oesophageal adenocarcinoma.

**Figure 3.**
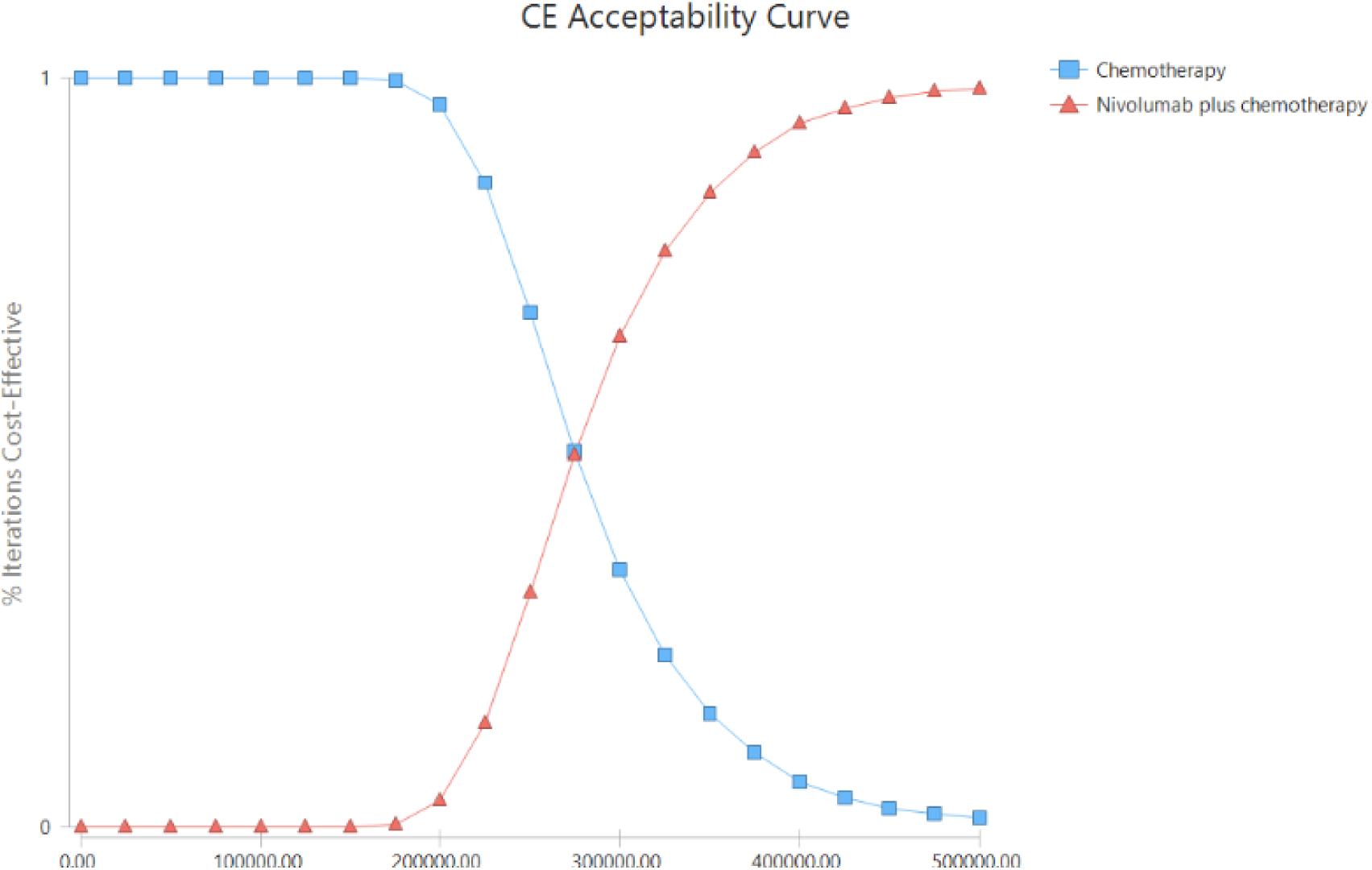
Acceptability Curves for the choice of sintilimab plus IBI305 versus sorafenib at different WTP thresholds in patients in the treatment of hepatocellular carcinoma. Abbreviation: WTP= Willingness to pay.

**Figure 4.**
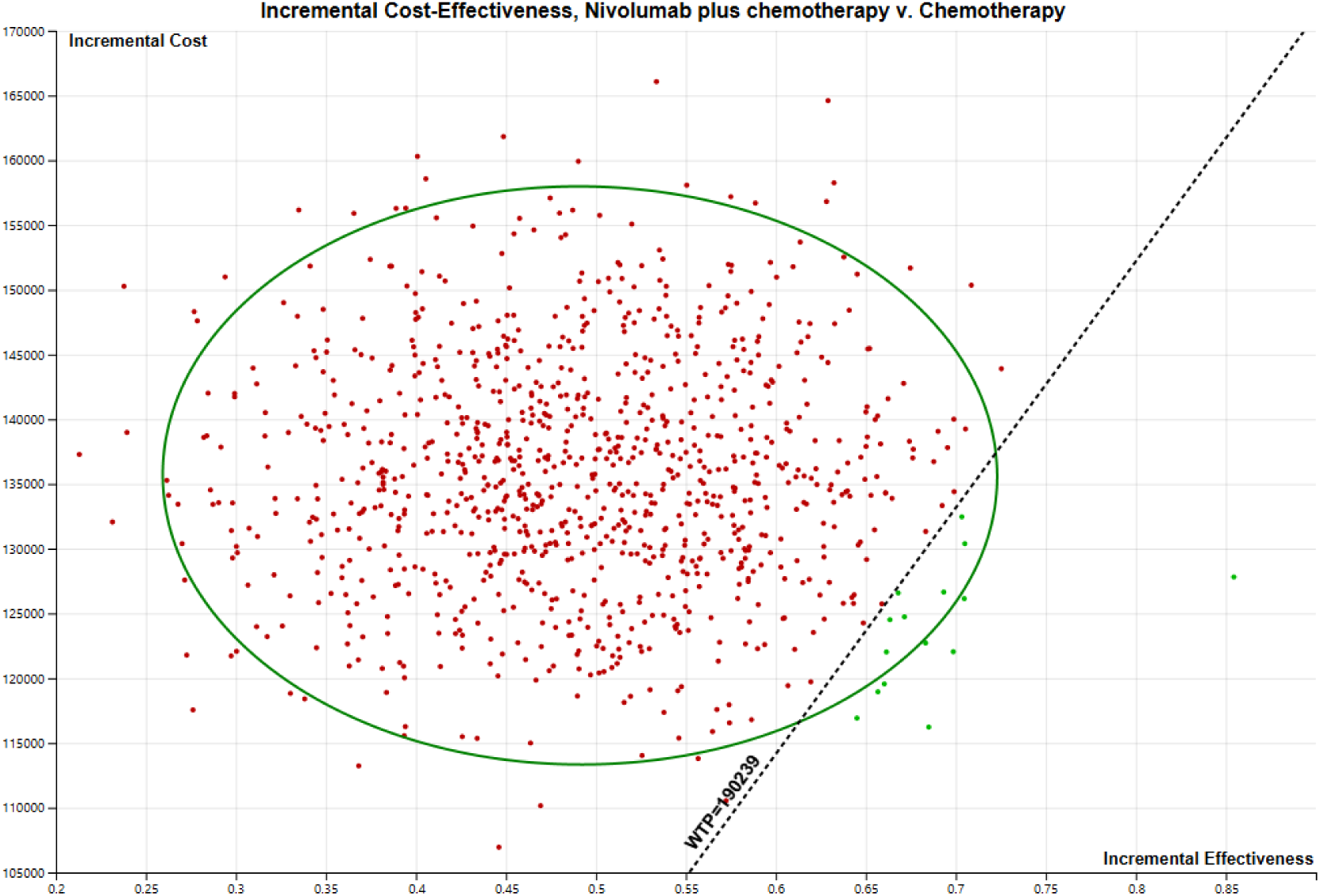
Incremental cost-effectiveness scatterplots.

We also conducted subgroup analysis of the patients with PD-L1 CPS≥5 according to Median age (IQR),Race(Asian or Non-Asian),Region (Asia or United States and Canada or Rest of world),Primary tumour location at initial diagnosis (GC or GEJC or EAC), the value of Tumour cell PD-L1 expression, Eastern Cooperative Oncology Group performance status(0 or 1),Prior surgery(yes or no), the number of Organs with metastases, Signet ring cell carcinoma(yes or no), Lauren classification (Intestinal type or Diffuse type or Mixed or Unknown), MSI status (MSS or MSI-H or Not reported or invalid),Chemotherapy regimen(FOLFOX or XELOX),Disease stage (Metastatic or Locally advanced or Locally recurrent), Site of metastases (Liver Or Peritoneum or CNS) according to the survival of subgroups of patients in CheckMate 649. Unfortunately,all subgroup analysis factors are all not likely to be cost effective when the payment threshold Less than or equal to $207659 per QALY.

## 4. Discussion

Fluoropyrimidine plus platinum-based chemotherapy is a first-line treatment for unresectable advanced or metastatic human epidermal growth factor receptor 2 (HER2) negative gastric and GEJ adenocarcinoma, but its efficacy is poor. The study subjects included in CheckMate 649 are patients with advanced/metastatic gastric cancer/gastroesophageal junction cancer/oesophageal adenocarcinoma with negative HER2. Nivolumab plus chemotherapy was compared with chemotherapy to evaluate whether Nivolumab plus chemotherapy could be used as first-line therapy for gastric/GEJ/oesophageal adenocarcinoma patients with negative HER2 .The result of CheckMate 649 showed that Nivolumab plus chemotherapy had significantly improved median survival time and median progression-free survival time compared to chemotherapy.Although Nivolumab plus chemotherapy has considerable efficacy in patients with gastric/GEJ/oesophageal adenocarcinoma, economic factors cannot be ignored. Many cancer patients are often forced to ignore the most effective drugs for their treatment because of high drug prices and treatment costs. Therefore, in order to avoid wasting medical resources, it is necessary to conduct a cost-benefit analysis.

Based on CheckMate 649 clinical trials and the latest population data and drug prices in USA, Our study is the first to evaluate whether Nivolumab plus chemotherapy is more cost-effective than chemotherapy as first-line treatment for patients with gastric/GEJ/oesophageal adenocarcinoma from the American payers’ perspectives.We obtained the data of Nivolumab plus chemotherapy group and chemotherapy group through the clinical trial CheckMate 649, so we could only conduct cost-effectiveness analysis on these two groups.Our analysis shows that with a WTP threshold of $207659/QALY for the two groups,the Nivolumab plus chemotherapy group may not be cost-effective to be the first-line for gastric/GEJ/oesophageal adenocarcinoma. We also performed subgroup analyses for all the subgroups mentioned in the clinical trials, unfortunately, the subgroup analysis implied that Nivolumab plus chemotherapy was not a cost-effective strategy across all the patient subgroups.

One-way sensitivity analysis demonstrated that reducing the cost of drugs was the most influential factor to the result of cost-effectiveness. Since the therapeutic effect of the experimental group was better than that of the control group, and chemotherapy was in the control group, the price largely determines the cost-effectiveness. The lower the price of capecitabine, oxaliplatin, leucovorin and fluorouracil, the lower the cost-effectiveness of the experimental group. There also have been some economic studies on chemotherapy for gastric/GEJ/oesophageal adenocarcinoma, but mainly compared with chemotherapy regimens such as FOLFOX and XELOX. Chemotherapy is much less expensive than immunotherapy in most cases, so it is easier to show cost-effectiveness. Another cost-benefit analysis of Atezolizumab and bevacizumab combination compared with sorafenib in unresectable hepatocellular carcinoma also showed that the immunotherapy group was not cost-effective compared to sorafenib alone(28). Therefore, We conclude that PD-1 inhibitors may be difficult to recommend as cost-effectiveness options for first-line recommendations in advanced gastric/GEJ/oesophageal adenocarcinoma. We found that when the cost of Nivolumab plus chemotherapy was reduced by 23%,respectively, the ICER was $207659 /QALY which was cost-effective. Therefore, changing the price of Nivolumab plus chemotherapy is an effective feasible strategy to achieve efficient use of them. medical insurance authority could negotiate with pharmaceutical companies to ensure reasonable drug prices and adjust the medical insurance list to reduce the medical burden on patients.

## 5. Limitations

Our study has some limitations. First, CheckMate 649 is a phase three randomised controlled trial, and we used this model to simplify the study. For instance, regarding the adverse reactions, we selected the three to four main AEs that would lead to errors. Second, the study’s data originated from the CheckMate 649 trial. Due to the limitation of the number of patients included in the trial, we could not perform a larger-scale analysis, and the trial did not provide the follow-up survival data for patients. We relied on the survival data based on the trial and performed a reasonable extrapolation to predict the long-term survival of patients. This will inevitably be different from the data of real-world patients obtained through regular follow-ups. Third, since CheckMate 649 does not disclose the specific health data of patients, our PFS and PD effectiveness were derived from previously published related studies. This may be different from the actual situation of the study. Fourth, we only considered the cost impact and utility reduction caused by the three to four main AEs. The utility reduction caused by specific adverse reactions comes from other published literature, which is in line with the real situation. Fifth, the clinical trial is a multicentred and comprehensive study between different countries and races. The treatment plan of the trial, and especially the follow-up treatment of patients, will be adjusted appropriately according to the specific situation.Therefore, more clinical trials are still required to reduce the study population, follow-up treatment, and other factors that impact the results.

## 6. Conclusion

Overall, from the Americans payers’ perspectives, compared with chemotherapy, Nivolumab plus chemotherapy a for the first-line treatment of patients aged 18 years or older who were clinically or pathologically diagnosed with locally advanced or metastatic gastric/GEJ/oesophageal adenocarcinoma may be not a cost-effective choice at a WTP threshold of $207659 /QALY.

## Data Availability

All relevant data are within the manuscript and its Supporting Information files

## Acknowledgements

The authors wish to acknowledge Dr Xiang-ping Li, Professor of Department of Pharmacy, Xiangya Hospital, Central South University, for his help in tnterpreting the significance of the results of the study.And also acknowledge Dr Qin Zhou, Professor of Department of Oncology, Xiangya Hospital, Central South University, for support funding to this study.

## Ethics approval

This study was based on a literature review and modelling techniques; this study did not require approval by an institutional research ethics board.

## Disclaimer

The views expressed are those of the authors. The funding agencies played no role in the study design, data collection and analysis, decision to publish, or manuscript preparation.

## Provenance and peer review

Not commissioned; externally peer reviewed.

## Data sharing statement

No additional data are available.

## Supplemental material

Supplemental material for this article is available online.

## Ethics approval and consent to participate

Not applicable

## Consent for publication

Not applicable

## Availability of data and materials

Data sharing is not applicable to this article as no datasets were generated or analysed during the current study.

## Authors’ contributors

Mr Zhou and Mr Tang had full access to all of the data in the study and takes responsibility for the integrity of the data and the accuracy of the data analysis.

Concept and design: Li.

Acquisition, analysis, or interpretation of data: Zhou,Tang. Drafting of the manuscript: Zhou,Tang,Li.

Critical revision of the manuscript for important intellectual content: Tang. Statistical analysis: Zhou,Tang.

Obtained funding: Zhou.

Administrative, technical, or material support: Zhou,Tang. Supervision:Tang.

